# An Analysis of Controlled Human Infection Studies Registered on ClinicalTrials.gov

**DOI:** 10.1101/2024.03.13.24304104

**Authors:** Danny Toomey, Jupiter Adams-Phipps, James Wilkinson, John Pietro, Jake Littman, Steffen Kamenicek, Daniel Kaufman, Euzebiusz Jamrozik, Joshua Osowicki, Meta Roestenberg, Ian J. Saldanha

## Abstract

**OBJECTIVES:** Controlled human infection studies (CHIS) involve intentional exposure of human volunteers to infectious agents. Given the small size of CHIS, aggregating data across studies is critical to the field. The objectives of the current analysis were to (1) evaluate the use of ClinicalTrials.gov for CHIS data reporting and (2) compare CHIS clinical trial participant flow and AE reporting in ClinicalTrials.gov with the same data in corresponding published articles.

**DESIGN:** ClinicalTrials.gov records that described a CHIS were included and data were accessed using the AACT API. These data were compared with results extracted from publications associated with included records’ NCT identifiers and compared in groups stratified by sponsor type, cohort size, and risk of bias in selection of the reported result as determined by Domain 5 of the Cochrane Risk of Bias 2.0 tool.

**RESULTS:** We screened 5,131 ClinicalTrials.gov records for inclusion, reviewed 410 records in full, and included 344 records. The overall prevalence of any discrepancy was 40%. Compared with their respective groups, significant discrepancies were observed in publicly funded trials, trials in the 3rd quartile of study size, and trials with a high risk of bias in selection of the reported result. Five studies reported a total of five SAEs in ClinicalTrials.gov records but not in any associated publications.

**CONCLUSION:** We identified an overall prevalence of discrepancy of 40% in CHIS, which is comparable to the prevalence observed in other types of clinical trials. In general, medium-sized, publicly funded trials tended to have more discrepancies in reporting, which may reflect the resources typically available to large, privately funded trials or the relative ease of reporting in smaller trials with fewer overall AEs. These results highlight the need to facilitate clear and consistent reporting in CHIS.

**Strength and Limitations:**

- This is the first study comparing CHIS AE reporting with trial registry data.
- This study contributes to a sparse general literature on reporting discrepancies.
- We provide recommendations for best practices to reduce the problems we identify.
- Our data likely exhibit heterogeneity arising from our aggregating across studies of different infectious agents.
- Only ClinicalTrials.gov was evaluated, and it is possible that trials that would have met inclusion criteria are registered in other databases.

## 1 Introduction

Controlled human infection studies (CHIS) model an encounter between human hosts and pathogens by deliberately exposing selected volunteers to a well-characterized infectious agent under controlled conditions [1,2]. CHIS are used for many purposes, such as studying the transmission and pathogenesis of infectious diseases and evaluating the efficacy of vaccines or other interventions [3]. Records of such trials date back to the 18^th^ century, although many early challenge experiments would not have met modern ethical research standards set forth in the 1970s [4]. Although CHIS are a powerful tool that can be used to expedite the development of vaccines and therapies for infectious diseases [1,5–7], their use has been relatively sporadic, potentially reflecting ethical or efficacy concerns or a lack of sustained investment [3,4,8]. The benefits of the data gathered in CHIS would be enhanced by comprehensive reporting as well as standardization of study protocols. Additionally, the use of data sharing principles such as Findability, Accessibility, Interoperability, and Reuse (FAIR) greatly aid in the aggregation and reuse of CHIS data, which is vital due to their small size compared to other clinical trials [9].

To investigate the safety of modern CHIS, we previously conducted a systematic review of adverse events (AEs) and serious adverse events (SAEs) in CHIS published between 1980 and 2021 [10]. Serious adverse events (SAEs) are defined as an adverse event that results in a serious outcome, such as hospitalization, permanent disability, and death [11]. Some AEs that occur during a study may be directly related to study interventions while others may be incidental. Our previous review found that a minority of participants in modern CHIS experienced challenge-related severe AEs (as defined by study authors) or SAEs [10]. Across 187 studies that reported SAE data, 23 of 10,016 participants (0.2%) experienced at least one SAE. The most frequent SAEs were severe vomiting and/or diarrhea, hepatitis, and hyperbilirubinemia. Across 94 studies that reported data on severe AEs (grade 3 or higher), between 285 and 801 of 5,083 participants^1^ (5.6% to 15.8%) experienced at least one severe AE [10].

Although the above findings generally support the safety of modern CHIS, the review also identified issues related to non-standardized reporting of CHIS [10,12], including regarding the classification of this type of research. Previous work has discussed the wide ranging terminology in use for CHIS [12], with the predominant issues posed being difficulty identifying CHIS across different models and fields. This ambiguity is related to a lack of consistency with what constitutes a ‘challenge’ with a microorganism. Some intuitive definitions, such as defining a challenge agent as a known infectious organism, would exclude studies evaluating therapeutic infection or phase 1 live attenuated vaccine trials. Studies aiming to produce colonization rather than symptomatic infection are another borderline case. The review also found an inconsistent use of trial registries [10], with approximately 75% of CHIS started in the 2010’s listed in at least one registry.

ClinicalTrials.gov, the world’s largest repository for clinical trial data, was launched in February 2000 by the US National Institutes of Health (NIH) and the FDA as a public registry of medical studies in human volunteers, and is maintained by the US National Library of Medicine [13–15]. Database fields detailing study AEs were subsequently made publicly available in September 2008 [13]. Although the rate of study registration has increased over time as a result of regulatory requirements and voluntary registration on the part of sponsors and investigators [13], some studies still do not get registered on ClinicalTrials.gov or any other registry. Among those registered, many fail to post results, with a recent report identifying over 3,000 clinical trials across all fields with missing results that have been overdue since February 2018 [16].

A key finding of our previous review was that AE reporting is often unclear or missing in CHIS publications [10]. To understand whether AE reporting is more clear or complete in ClinicalTrials.gov records than in publications, the current analysis (1) evaluates the use of ClinicalTrials.gov for CHIS registration and data reporting and (2) compares CHIS clinical trial participant flow and AE reporting in ClinicalTrials.gov with the same data in corresponding published articles.

## 2 Methods

A full protocol is available in the supplementary materials. Patients and members of the public were not involved in the design of the study. This analysis was pre-registered at Prospero, number CRD42022330047.

### 2.1 Eligibility criteria for CHIS

Studies registered on ClinicalTrials.gov that involved intentional exposure of human volunteers to an infectious agent for the purpose of developing or using a model of infection, commonly known as CHIS or human challenge trials, were included. There is ongoing debate regarding the precise definition of a CHIS [12,17]. For our review, we examined studies that involved intentional exposure of human volunteers to wild-type or attenuated organisms (infectious agents). Challenges with candidate vaccine viruses were also included, as were studies in which previously challenged participants were challenged again with the same infectious agent (i.e., rechallenges). Studies involving live attenuated vaccines were only included when the vaccine strain was used as a challenge agent.

### 2.2 Identification of CHIS registered on ClinicalTrials.gov

Searches were performed as structured query language (SQL) queries using the Aggregated Analysis of ClinicalTrials.gov application programming interface (AACT API) on June 30, 2022. The search was updated on April 15, 2023. The full search strategy, including specific queries, is included in the Supplementary Materials. This search was conducted as a systematic review according to PRISMA guidelines [18].

### 2.3 Identification of Full Publications of CHIS

We used two strategies to identify full publications of CHIS: (1) publications listed in the ClinicalTrials.gov record for each included CHIS, and (2) to identify additional articles that were not listed in the ClinicalTrials.gov record, a PubMed search using each National Clinical Trial (NCT) number. Articles were included if they were peer-reviewed and reported AEs in the CHIS linked by NCT number. Articles published before the date the record was first posted on ClinicalTrials.gov were excluded, as were articles reporting secondary analyses of data from registered CHIS.

### 2.4 Study Categorization

Studies were categorized according to recruitment status to differentiate between those that were still recruiting, ongoing, suspended and/or terminated, withdrawn, completed, or of unknown status (missing updates). Studies that had been completed were further grouped by whether they posted results on ClinicalTrials.gov and whether a corresponding published article discussing results of the study was listed within the ClinicalTrials.gov study record or identifiable through a PubMed search for the record’s NCT number.

### 2.5 Screening and Data Extraction Process

Each record was screened independently by two of six investigators. Titles and study descriptions were reviewed to evaluate whether the record described a CHIS. Where feasible, data were automatically extracted from the AACT database. Data that were extracted automatically were independently reviewed by at least two reviewers for verification. Data that could not be extracted automatically were extracted manually by two reviewers working independently. For records with a corresponding published article discussing results from the same study, data were likewise extracted manually. Any discrepancies in screening or data extraction were either resolved by discussion among the reviewers or by JAP or DT. Participant flow and AE data were not extracted from associated publications if results were not posted on the ClinicalTrials.gov record, as data from both a ClinicalTrials.gov record and a publication are necessary for comparison. Studies registered on ClinicalTrials.gov less than 1 year before April 15, 2023 (the date of our last query of the database) were excluded.

### 2.6 Assessment of risk of bias in the selection of the reported result

Risk of bias in the selection of the reported result was assessed using domain 5 of the Cochrane Risk of Bias 2.0 Tool [19]. Assessments were performed by DT and confirmed by 1 of 6 reviewers. Disputes were resolved by JAP or DT.

### 2.7 Synthesis Methods

ClinicalTrials.gov records that posted results and included at least one publication were included in data synthesis. Data were tabulated to create summary statistics by relevant parameters. We calculated the prevalence of discrepancies between reporting in ClinicalTrials.gov records and associated publications. Data were analyzed by study sponsor type (private, public, and public-private partnership), cohort size (1^st^, 2^nd^, 3^rd^, and 4^th^ quartiles), and risk of bias in the selection of the reported result (low, some concerns, and high). Where applicable, data for the number of volunteers challenged, infected, with AEs, or with SAEs were recorded as a range to account for ambiguous reporting.

Studies sponsored by government organizations were categorized as public, studies sponsored by private for-profit or not-for-profit organizations were categorized as private, and studies sponsored by an organization that was formed as an independent collaborative partnership between a governmental organization and a private organization were categorized as public-private partnerships.

Discrepancies were defined as any instance in which the same metric (number challenged, infected, with AEs, or with SAEs) was unambiguously reported differently in the record and its associated publications. Odds ratios (ORs) and 95% confidence intervals (CIs) were used to compare the likelihoods of discrepancy. These analyses were performed *post hoc*. Records with data recorded as a range due to ambiguous reporting were excluded from statistical analysis. The relationship between study size, risk of bias, and sponsor type was evaluated *post hoc* using one-way ANOVAs for continuous data and chi-square tests of independence for categorical data. Statistical significance was defined at a 5% level for all analyses.

## 3 Results

### 3.1 Study Selection

The search returned 5,131 records, of which 4,721 records were excluded during screening (Supplementary Figure 1). The remaining 410 records were screened in full, of which 66 were excluded for not meeting the definition of a CHIS. The remaining 344 records met inclusion criteria. A complete list of records excluded for these reasons is included in the Supplementary Materials.

### 3.2 Reporting in Individual CHIS

Among the 344 included CHIS, 264 (76.7%) were completed, 13 (3.8%) were active and not recruiting, 22 (6.4%) were either recruiting or enrolling by invitation, 12 (3.5%) were terminated, and 16 (4.7%) were of unknown status. Among all 264 completed CHIS, 66 (25.0%) posted results, and 156 (59.1%) listed at least one published article. Among the 344 included CHIS, there were 46 studies (13.4%) that both posted results and linked to a publication, and 209 studies (60.8%) that had results available in some form (either as results posted on ClinicalTrials.gov or by linking to a publication) (Supplemental Table 1).

### 3.3 Challenges, Infections, and Adverse Event Reporting

Among completed CHIS, 46 ClinicalTrials.gov records both reported results and listed at least one associated publication, with 195 associated publications listed in total (median 1, range 1 to 82 publications per CHIS). The most common challenge agents were plasmodium species (17 records), influenza (8 records), respiratory syncytial virus (4 records), and rhinovirus (4 records). Data were not extracted from the 474 associated publications whose ClinicalTrials.gov record did not post results. Where applicable, data are provided as ranges to account for ambiguous reporting. A total of 3,574 volunteers were enrolled, of whom between 2,998 (83.9%) and 3,131 (87.6%) were challenged with a pathogen, and between 1,297 (41.4 - 43.3%) and 1,608 (51.4-53.6 %) were reported to have laboratory confirmed infection or symptoms diagnostic for infection. In associated publications, total reported enrollment was 3,399, with 2,615 (76.9%) to 2,620 (77.1%) challenged volunteers and 1,437 (54.8 - 54.9%) to 1,455 (55.5 - 55.6%) confirmed infections. ClinicalTrials.gov records reported that 1,921 (61.4 - 64.1%) to 2,166 (69.2 - 72.2%) volunteers experienced at least one AE, whereas associated publications reported 1,532 (58.5 - 58.6%) to 1,730 (66.0 - 66.2%) volunteers experienced at least one AEs. Twenty-nine SAEs were reported in ClinicalTrials.gov records and 25 SAEs were reported in associated publications (Table 1).

**Table 1.**
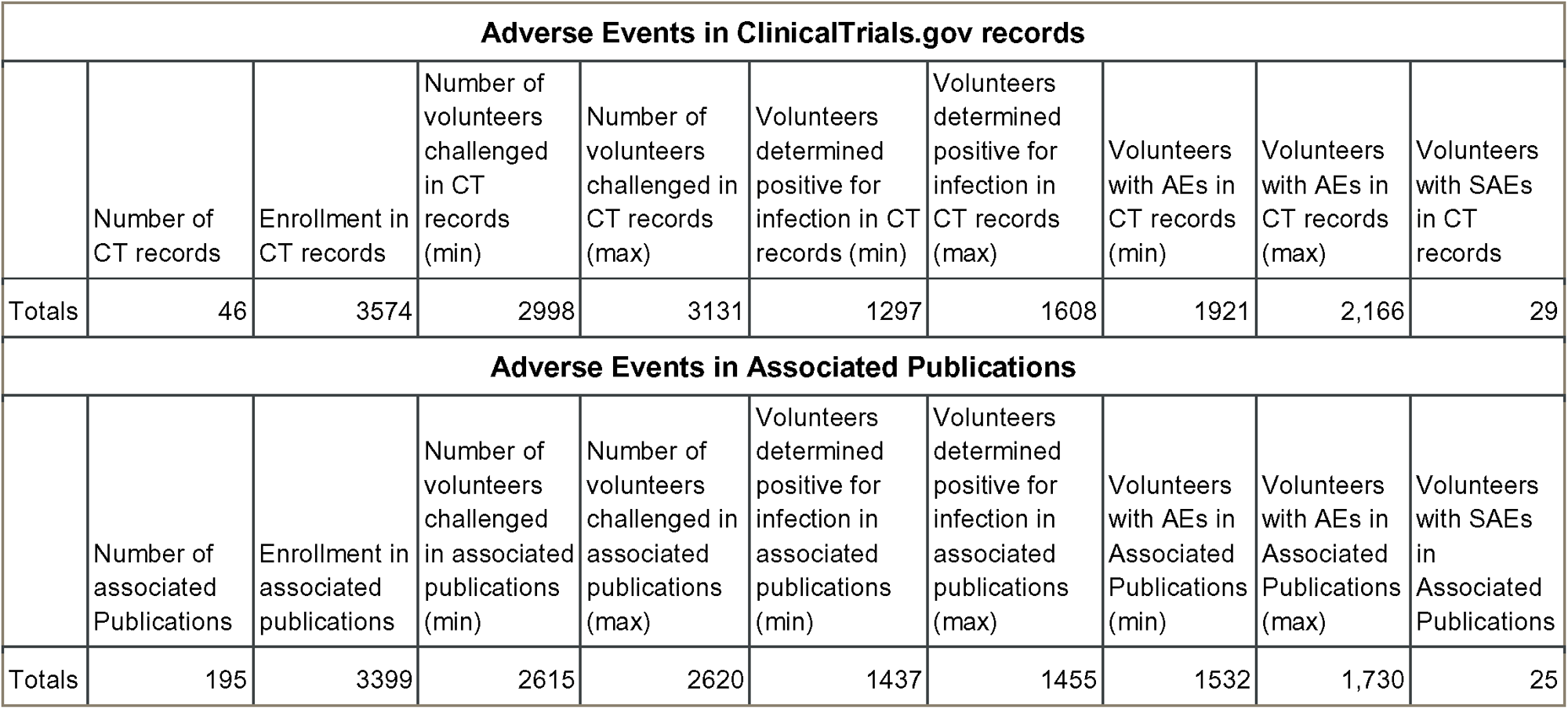
Adverse Events in ClinicalTrials.gov records and Associated Publications.

### 3.4 Comparison of Results Reported in ClinicalTrials.gov Records versus Associated Publications

Completed records that both posted results to ClinicalTrials.gov and linked to a published article discussing results were compared to identify discrepancies in data reporting. Among included records, 23 were sponsored by private organizations, 18 by public organizations, and 5 by public-private partnerships, with 60, 127, and 8 associated publications, respectively (Table 2). Results were posted for 25.7%, 14.6%, and 50.0% of privately sponsored records, publicly sponsored records, and public-private partnerships, respectively. Records were divided into quartiles by the number of volunteers enrolled, with the 1st, 2nd, 3rd, and 4th quartiles being 6 to 26 volunteers, 27 to 58 volunteers, 59 to 79 volunteers, and 80 to 440 volunteers, respectively (Table 3). Twenty-six ClinicalTrial.gov records had low risk, 1 had some concern, and 19 had high risk of bias related to selection of the reported result (Table 4).

**Table 2.** Adverse Events by Sponsor in ClinicalTrials.gov Records and Associated Publications.

| **ClinicalTrials.gov Adverse Events by Sponsor** |  |  |  |  |  |  |  |  |  |
| --- | --- | --- | --- | --- | --- | --- | --- | --- | --- |
| Sponsor Type | Number of CT records included in analysis | Enrollment in CT records | Number of volunteers challenged in CT records (min) | Number of volunteers challenged in CT records (max) | Volunteers determined positive for infection in CT records (min) | Volunteers determined positive for infection in CT records (max) | Volunteers with AEs in CT records (min) | Volunteers with AEs in CT records (max) | Volunteers with SAEs in CT records |
| Private | 23 | 2337 | 2050 | 2062 | 786 | 870 | 1298 | 1541 | 18 |
| Public | 18 | 1165 | 879 | 998 | 483 | 710 | 560 | 562 | 10 |
| Public-Private | 5 | 72 | 69 | 71 | 28 | 28 | 63 | 63 | 1 |
| *Totals* | *46* | *3574* | *2998* | *3131* | *1297* | *1608* | *1921* | *2166* | *29* |
| **Associated Publications Adverse Events by Sponsor** |  |  |  |  |  |  |  |  |  |
| Sponsor Type | Number of associated Publications | Enrollment in associated publications | Number of volunteers challenged in associated publications (min) | Number of volunteers challenged in associated publications (max) | Volunteers determined positive for infection in associated publications (min) | Volunteers determined positive for infection in associated publications (max) | Volunteers with AEs in Associated Publications (min) | Volunteers with AEs in Associated Publications (max) | Volunteers with SAEs in Associated Publications |
| Private | 60 | 2238 | 1757 | 1760 | 785 | 800 | 1167 | 1293 | 15 |
| Public | 127 | 1089 | 789 | 789 | 598 | 599 | 301 | 372 | 9 |
| Public-Private | 8 | 72 | 69 | 71 | 54 | 56 | 64 | 65 | 1 |
| *Totals* | *195* | *3399* | *2615* | *2620* | *1437* | *1455* | *1532* | *1730* | *25* |

**Table 3.** Adverse Events by Study Size in ClinicalTrials.gov and Associated Publication.

**ClinicalTrials.gov Adverse Events by Study Size**
| Study Size | Number of CT records included in analysis | Enrollment in CT records | Number of volunteers challenged in CT records (min) | Number of volunteers challenged in CT records (max) | Volunteers positive for infection in CT records (min) | Volunteers positive for infection in CT records (max) | Volunteers with AEs in CT records (min) | Volunteers with AEs in CT records (max) | Volunteers with SAEs in CT records |
| --- | --- | --- | --- | --- | --- | --- | --- | --- | --- |
| 1st Quartile - 6 to 26 volunteers | 12 | 194 | 178 | 186 | 106 | 106 | 121 | 121 | 2 |
| 2nd Quartile - 27 to 58 volunteers | 11 | 493 | 456 | 461 | 226 | 252 | 321 | 341 | 6 |
| 3rd Quartile - 59 to 79 volunteers | 11 | 721 | 660 | 670 | 339 | 383 | 486 | 503 | 6 |
| 4th Quartile - 80 to 440 volunteers | 12 | 2166 | 1704 | 1814 | 626 | 867 | 993 | 1201 | 15 |
| *Totals* | *46* | *3574* | *2998* | *3131* | *1297* | *1608* | *1921* | *2166* | *29* |

**Associated Publication Adverse Events by Study Size**
| Study Size | Number of associated Publications | Enrollment in associated publications | Number of volunteers challenged in associated publications (min) | Number of volunteers challenged in associated publications (max) | Volunteers positive for infection in associated publications (min) | Volunteers positive for infection in associated publications (max) | Volunteers with AEs in Associated Publications (min) | Volunteers with AEs in Associated Publications (max) | Volunteers with SAEs in Associated Publications |
| --- | --- | --- | --- | --- | --- | --- | --- | --- | --- |
| 1st Quartile - 6 to 26 volunteers | 100 | 140 | 137 | 139 | 112 | 115 | 107 | 112 | 1 |
| 2nd Quartile - 27 to 58 volunteers | 40 | 364 | 341 | 341 | 268 | 268 | 222 | 267 | 4 |
| 3rd Quartile - 59 to 79 volunteers | 28 | 687 | 492 | 492 | 320 | 332 | 348 | 324 | 7 |
| 4th Quartile - 80 to 440 volunteers | 27 | 2208 | 1645 | 1648 | 737 | 740 | 855 | 1027 | 13 |
| *Totals* | *195* | *3399* | *2615* | *2620* | *1437* | *1455* | *1532* | *1730* | *25* |

**Table 4.**
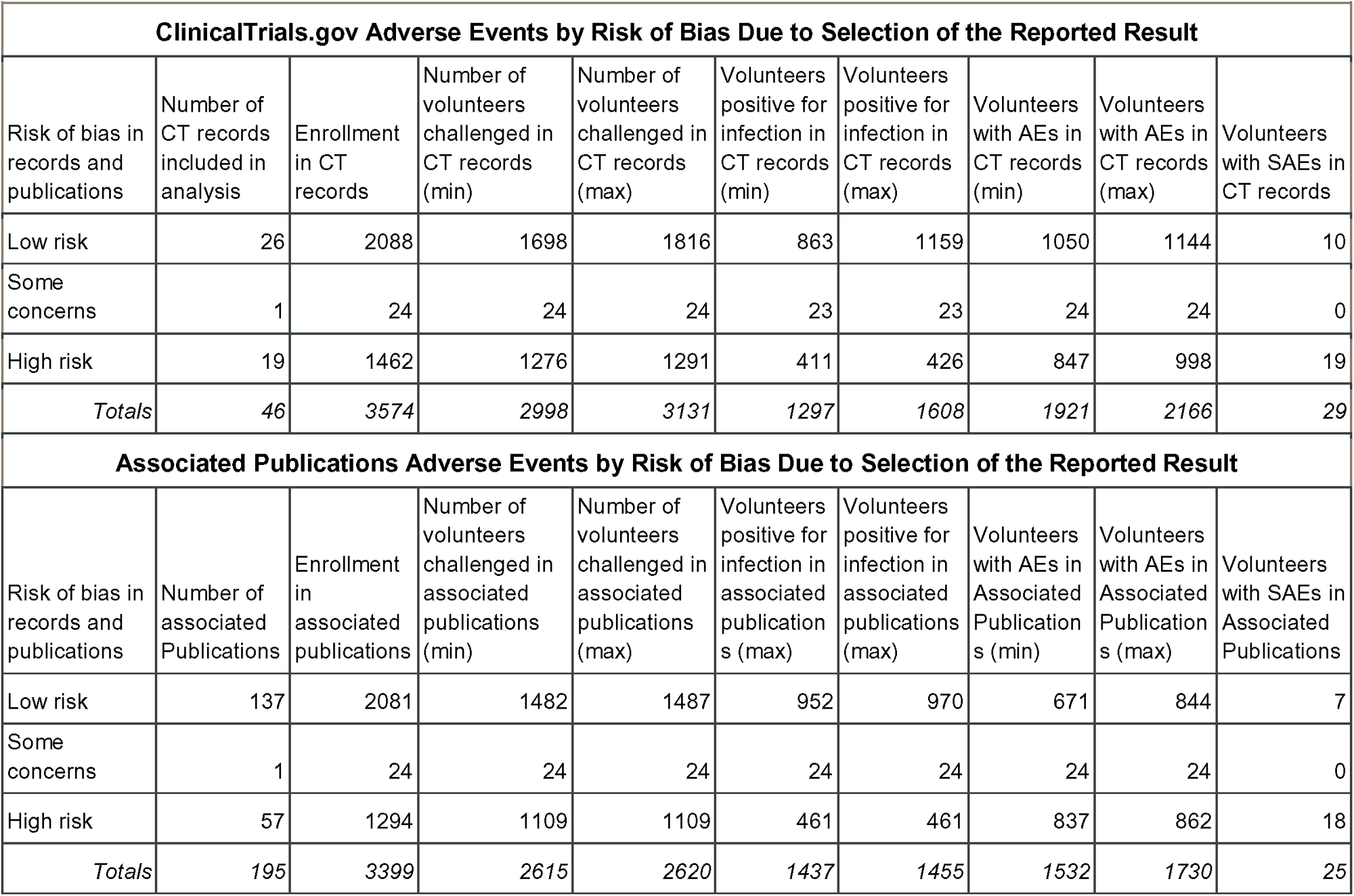
Adverse Events by Risk of Bias in ClinicalTrials.gov Records and Associated Publications.

| **ClinicalTrials.gov Adverse Events by Risk of Bias Due to Selection of the Reported Result** |  |  |  |  |  |  |  |  |  |
| --- | --- | --- | --- | --- | --- | --- | --- | --- | --- |
| Risk of bias in records and publications | Number of CT records included in analysis | Enrollment in CT records | Number of volunteers challenged in CT records (min) | Number of volunteers challenged in CT records (max) | Volunteers positive for infection in CT records (min) | Volunteers positive for infection in CT records (max) | Volunteers with AEs in CT records (min) | Volunteers with AEs in CT records (max) | Volunteers with SAEs in CT records |
| Low risk | 26 | 2088 | 1698 | 1816 | 863 | 1159 | 1050 | 1144 | 10 |
| Some concerns | 1 | 24 | 24 | 24 | 23 | 23 | 24 | 24 | 0 |
| High risk | 19 | 1462 | 1276 | 1291 | 411 | 426 | 847 | 998 | 19 |
| *Totals* | *46* | *3574* | *2998* | *3131* | *1297* | *1608* | *1921* | *2166* | *29* |
| **Associated Publications Adverse Events by Risk of Bias Due to Selection of the Reported Result** |  |  |  |  |  |  |  |  |  |
| Risk of bias in records and publications | Number of associated Publications | Enrollment in associated publications | Number of volunteers challenged in associated publications (min) | Number of volunteers challenged in associated publications (max) | Volunteers positive for infection in associated publications (max) | Volunteers positive for infection in associated publications (max) | Volunteers with AEs in Associated Publications (min) | Volunteers with AEs in Associated Publications (max) | Volunteers with SAEs in Associated Publications |
| Low risk | 137 | 2081 | 1482 | 1487 | 952 | 970 | 671 | 844 | 7 |
| Some concerns | 1 | 24 | 24 | 24 | 24 | 24 | 24 | 24 | 0 |
| High risk | 57 | 1294 | 1109 | 1109 | 461 | 461 | 837 | 862 | 18 |
| *Totals* | *195* | *3399* | *2615* | *2620* | *1437* | *1455* | *1532* | *1730* | *25* |

#### a) Relationships Between Variables

To evaluate the relationship between sponsor type, risk of bias in selection of the reported result, and study cohort size, several *post hoc* analyses were performed. The relationship between sponsor type and study size was evaluated by a one-way ANOVA, which did not indicate a significant relationship (F=2.62, df=2, p=0.08). The relationship between risk of bias in selection of the reported result and study size was evaluated by a one-way ANOVA, which did not indicate a significant relationship (F=0.20, df=2, p=0.82). The relationship between sponsor type and risk of bias in selection of the reported result was evaluated by a chi-square test for independence, which did not indicate a significant relationship (X=0.69, df=8, p=0.99).

#### b) Volunteers Challenged

Twenty-eight records were included in statistical analysis for discrepancies among the number of volunteers challenged, with discrepancies observed in 14.3%. Publicly sponsored studies were more likely than other studies to have discrepancies (OR: 2.43, 95% CI: 1.10-5.37). Studies in the 2nd quartile (OR: 3.00, 95% CI: 1.35-6.64) and 3rd quartile (OR: 5.00, 95% CI: 2.25-11.11) of study size were more likely than studies in the other quartiles to have discrepancies. Studies with a low risk of bias in selection of the reported result were more likely than studies with some concerns or studies with a high risk of bias to have discrepancies (OR: 3.00, 95% CI: 1.24-7.25), and studies with a high risk of bias were less likely than studies with some concerns or a low risk of bias to have discrepancies (OR: 0.39, 95% CI: 0.17-0.94) (Table 5). Among the 4 records with discrepancies in the number of volunteers challenged, ClinicalTrials.gov records were more complete in all instances.

**Table 5.** Odds Ratio Comparisons.

|  | Challenged |  |  | Infected |  |  | with AEs |  |  | with SAEs |  |  |
| --- | --- | --- | --- | --- | --- | --- | --- | --- | --- | --- | --- | --- |
|  | OR | 95% CI LB | 95% CI UB | OR | 95% CI LB | 95% CI UB | OR | 95% CI LB | 95% CI UB | OR | 95% CI LB | 95% CI UB |
| *Sponsor type* |  |  |  |  |  |  |  |  |  |  |  |  |
| Privately vs not private | 0.85 | 0.38 | 1.87 | 1.45 | 0.64 | 3.33 | 0.80 | 0.46 | 1.40 | 0.41 | 0.15 | 1.17 |
| Public vs not public | 2.43* | 1.10* | 5.37* | 1.40 | 0.63 | 3.09 | 1.75 | 0.99 | 3.10 | 4.89* | 1.65* | 14.50* |
| Public-Private vs not public-private | 0.00 | N/A | N/A | 0.00 | N/A | N/A | 0.70 | 0.40 | 1.22 | 0.00 | N/A | N/A |
| *Study size* |  |  |  |  |  |  |  |  |  |  |  |  |
| 4th quartile vs not 4th | 0.00 | N/A | N/A | 0.00 | N/A | N/A | 0.30* | 0.17* | 0.54* | 0.00 | N/A | N/A |
| 3rd quartile vs not 3rd | 5.00* | 2.25* | 11.11* | 10.67* | 4.23* | 26.88* | N/A** | N/A | N/A | 1.44 | 0.55 | 3.73 |
| 2nd quartile vs not 2nd | 3.00* | 1.35* | 6.64* | 0.00 | N/A | N/A | 0.55* | 0.31* | 0.96* | 2.08 | 0.82 | 5.31 |
| 1st quartile vs not 1st | 0.00 | N/A | N/A | 1.40 | 0.63 | 3.09 | 0.18* | 0.10* | 0.33* | 1.44 | 0.55 | 3.73 |
| *Risk of bias* |  |  |  |  |  |  |  |  |  |  |  |  |
| Low vs not low | 3.00* | 1.24* | 7.26* | 0.56 | 0.24 | 1.29 | 0.80 | 0.46 | 1.40 | 0.36 | 0.13 | 1.03 |
| Some concern vs not some concern | 0.00 | N/A | N/A | N/A** | N/A | N/A | 0.00 | N/A | N/A | 0.00 | N/A | N/A |
| High vs not high | 0.39* | 0.17* | 0.94* | 0.45 | 0.19 | 1.05 | 2.00* | 1.13* | 3.54* | 3.17* | 1.10* | 9.14* |

Higher OR indicates a higher prevalence of discrepancy in the reference group.
\*Significant at an alpha level of 0.05
\*\*The prevalence of discrepancy in the reference groups in these comparisons was 100%, precluding OR analysis

#### c) Volunteers Infected

Twenty-two records were included in statistical analysis for discrepancies among the number of volunteers with signs of infection after challenge, with discrepancies observed in 13.6%. Studies in the 3rd quartile of study size were more likely than studies in the other quartiles to have discrepancies in the number of volunteers infected after challenge (OR: 10.67, 95% CI: 4.23-26.88). All records with some concerns for bias had discrepancies in the number of volunteers infected after challenge (Table 5). Of the 3 records with discrepancies in the number of volunteers infected, associated publications were more complete compared to ClinicalTrials.gov in all instances.

#### d) AE Reporting

Twenty-one records were included in statistical analysis for discrepancies among the number of volunteers with AEs, with discrepancies observed in 61.9%. All 11 studies in the 3rd quartile of study size had discrepancies in the number of volunteers with AEs. Studies with a high risk of bias in selection of the reported result were more likely than studies with a low risk of bias or some concerns to have discrepancies in the number of volunteers with AEs (OR: 2.00, 95% CI: 1.13-3.54). Studies in the 1st quartile (OR: 0.18, 95% CI: 0.10-0.33), 2nd quartile (OR: 0.55, 95% CI 0.31-0.96), and 4th quartile (OR: 0.30, 95% CI: 0.17-0.54) of study size were less likely than studies in the 3rd quartile to have discrepancies in the number of volunteers with AEs (Table 5). Of the 13 records with discrepancies in the number of volunteers with AEs, ClinicalTrials.gov records were more complete in 9 (69.2%) instances and associated publications were more complete in 4 (30.8%) instances.

#### e) SAE Reporting

Thirty-four records were included in statistical analysis for discrepancies among the number of volunteers with SAEs, with discrepancies observed in 8.8%. Studies sponsored by publicly funded organizations were more likely than other trials to have discrepancies in the number of reported SAEs (OR: 4.89, 95% CI: 1.65-14.50). Studies with a high risk of bias were more likely than studies with a low risk of bias or some concerns to have discrepancies (OR: 3.17, 95% CI: 1.10-9.14) (Table 5). Of the 3 records with discrepancies in the number of volunteers with SAEs, ClinicalTrials.gov records were more complete in 2 (66.7%) instances and associated publications were more complete in 1 (33.3%) instance.

#### f) Overall Completeness of AE reporting

Across 71 studies (66 completed, 5 terminated) that reported results on ClinicalTrials.gov and in associated publications, between 696 and 818 participants experienced AEs that were reported in ClinicalTrials.gov records but not in associated publications. Between 257 and 348 participants experienced AEs that were reported in associated publications but not in ClinicalTrials.gov records. The median time to post results to the ClinicalTrial.gov record was 1,382 days (interquartile range [IQR] 774 to 2,191). Ninety-four percent of ClinicalTrials.gov records reported AEs individually, rather than grouping related symptoms together into sets, compared with 60.9% of associated publications. There were 6 to 13 AEs that were reported after rechallenge in ClinicalTrials.gov records that were not reported in associated publications.

There were five participants that each experienced one SAE that was reported in ClinicalTrials.gov records but not in any associated publication: fractured wrist, breast cancer in situ, peripheral parasitemia, ruptured Achilles tendon, and pulmonary embolism. One record (NCT01024686) was terminated and so is not included in analyses, but is included in this summary for completeness. The relatedness of these SAEs to challenge was not discussed in their respective records. No participants experienced SAEs that were reported in associated publications but not in ClinicalTrials.gov records (Supplementary Table 2). An additional six participants experienced SAEs that were reported in ClinicalTrials.gov records that had no associated publications: asthma, rhabdomyolysis, acute coronary syndrome, acute psychosis, and fetal death.

## 4 Discussion

In this analysis, we compared results available in ClinicalTrials.gov records with results available in these records’ associated publications and quantified the prevalence of discrepancies between these sources. The likelihoods of discrepancies among the number of volunteers challenged, infected, with AEs, and with SAEs were 14.3%, 13.6%, 61.9%, and 8.8% respectively. Among all records eligible for analysis, 39.1% had some discrepancy. We also identified 5 SAEs that were reported in ClinicalTrials.gov records and not discussed in any publication of the results.

Publicly funded trials and trials with a high risk of bias related to selection of the reported result were most likely to have discrepancies in reported SAEs. Trials in the 3^rd^ quartile of study size and trials with a high risk of bias related to selection of the reported result were more likely to have discrepancies in reported AEs than studies in other study size quartiles and other risks of bias, respectively. There were no significant relationships identified between sponsor type, risk of bias, and study size in the occurrence of discrepancies. Data on the amount of funding trials received were not available, and it is possible that this would be a relevant factor. For example, if large, privately funded trials typically have more resources than smaller, publicly funded trials, they may have the budget to ensure proper reporting, which may have explained these results. The lower prevalence of discrepancies observed in smaller studies may reflect the relative ease of consistent reporting when the overall number of events to report is low. It is also possible that ClinicalTrials.gov reporting is more complete due to constraints on the sharing of data in the process of preparing manuscripts for peer-review and publication. At the same time, 59% of trials completed listed at least one publication in their record while only 25% had posted results, which may contextualize the relative ease of publishing a limited amount of relevant data in a peer-reviewed journal as opposed to a complete set of results in a clinical trial record.

Issues with data reporting in clinical trials are not isolated to CHIS, with a body of literature showing that reporting discrepancies are prevalent in other types of clinical trials. A recent review of trials registered on ClinicalTrials.gov in Canada found that 32% neither reported their results nor discussed them in publications [20]. A review of outcome-related discrepancies between registry entries and published reports in orthodontic RCTs identified discrepancies in 47% of publications [21], while a review of oncology trials identified a 63% discrepancy in secondary outcomes described in protocols compared with final results [22]. A random sample of 300 trials posted on ClinicalTrials.gov identified a discrepancy prevalence of 32% for SAEs in records compared with SAEs in publications of results [23]. A study investigating reporting discrepancies in a random sample of 110 phase 3 or 4 trials with results posted on ClinicalTrials.gov found that 20% of trials have inconsistencies in reported primary outcomes [24]. Our findings regarding discrepancies in the reporting of CHIS are therefore similarly observed in other types of clinical research.

We previously reported that adverse event reporting in CHIS is inconsistent [10]. Though this is not unique to CHIS, there is arguably a greater benefit to consistent reporting due to the increased value of data aggregation in a field of research where studies are typically small and where key benefits include providing results more quickly and with fewer volunteers compared to alternative trial designs (e.g., vaccine field trials). In particular, data on the numbers of volunteers who are challenged, become infected, and subsequently experienced AEs and/or SAEs must be reported clearly, especially if related to challenge or other study procedures. Consistent reporting is important for a wide range of clinical studies. The European Union’s Clinical Trials Information System (EU CTIS) clinical trial registry provides an example of a platform that facilitates consistent reporting by integrating trial registration with outcome reporting. CTIS registration requires expected AEs to be predefined. Clear reporting of which expected and unexpected AEs are noted for volunteers who do and do not receive a challenge organism would help to ensure accurate descriptions of what volunteers experience.

A key finding of the current study is that reporting is clearer in a database than in publications because it’s more comprehensive and easier to compare due to standardization. Repositories also facilitate finding and aggregating data for future studies. Our experience aggregating data for the current study highlights the ClinicalTrials.gov AACT API as a powerful tool that promotes efficient data sharing with minimal modifications. We provide in the supplementary methods an open-source program that was used to generate our dataset and can be used by others [25]. Our focus on ClinicalTrials.gov is not to imply that it should be the repository for all clinical trials but rather to highlight aspects of its reporting requirements. On the data entry side, it is relatively simple to add fields by arm for AE data. On the data reuse side, the AACT API is powerful and easy to use, with a well-documented schema to simplify finding a data field of interest. These functionalities of ClinicalTrials.gov provide a simple method for sharing AE data and could be used more widely. It serves as a strong example of a platform that fulfills FAIR data sharing principles. We recommend that CHIS researchers add data fields alongside the AE and SAE outcomes for the number of volunteers challenged and infected in each arm. We make these recommendations with the goal of a) ensuring CHIS fulfill FAIR data sharing practices to the fullest degree and b) maximizing the contribution each CHIS volunteer makes to science by facilitating the ease with which other researchers may access and find new insights from their data.

Our study has the following limitations: 1) not every relevant record may have been caught by the ClinicalTrials.gov search query used; 2) not every publication associated with each record may have been identified by NCT number; 3) our results may have been influenced by publication bias, which favors significant results; 4) our data likely exhibit heterogeneity arising from aggregation across different infectious agents; and 5) only ClinicalTrials.gov was evaluated, and it is possible that trials that would have met inclusion criteria are registered in other databases.

In conclusion, we found that 40% of CHIS had at least one discrepancy between results reported in ClinicalTrials.gov records and publications describing the same results. Rather than this being unique to CHIS, we note that similar levels of suboptimal reporting have been observed in other types of trials. Publicly funded, medium-sized trials were more likely to have discrepancies in reporting, which may reflect the resources typically available to large, privately funded trials or the relative ease of reporting in smaller trials. To address these issues, we propose minimal amendments to CHIS researchers’ data entry workflow in ClinicalTrials.gov to facilitate automatic aggregation of CHIS data via the AACT API and highlight the EU CTIS as an example of an integrated registration system that facilitates clear and consistent reporting. Due to the benefits of CHIS data being shared for aggregation, it is imperative that volunteer outcomes be described in publicly available repositories so that study data can be used as broadly and effectively as possible.

## Supporting information

Main Data Sheet

## Data Availability

All data and analyses performed are publicly available at https://github.com/1DaySooner/CTgovSystematicReview

## Author contributions

DT and JAP conceived the study, designed the study, and performed data collection, data analysis, and manuscript writing and review. JW, SK, JL, JP, and DK performed data pipeline optimization, data collection, data review, and contributed to manuscript writing. EJ, JO, MR, and IJS assisted in study design and provided expert review of the manuscript.

---

**Supplementary Figure 1.**
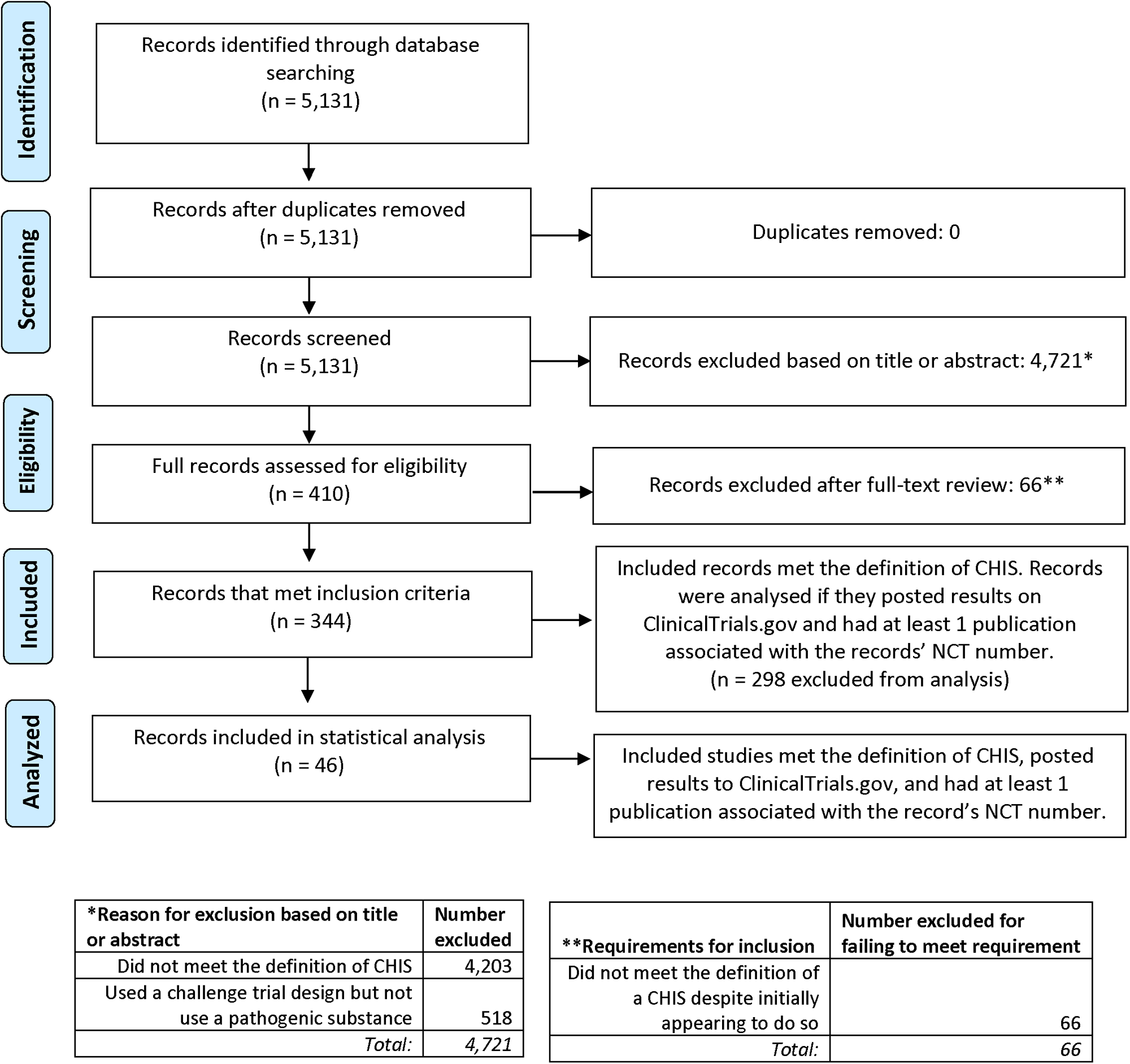
PRISMA Flowchart.

## ClinicalTrials.gov Systematic Review Protocol

### Objectives

The proposed review aims to investigate the use of ClinicalTrials.gov for CHIS registration and data reporting, and how this compares with data reporting in corresponding published articles describing CHIS. With the goal of evaluating the current use of ClinicalTrials.gov as a registration platform and recommending guidelines for future use, the present review seeks to answer the following research questions:

1. How many CHIS registered on ClinicalTrials.gov have at least one published article to report their results and how many post results to the CT record?

a. Reported in Section 3.2
2. Of those that post results, how many volunteers were challenged and infected? How many experienced AEs and SAEs?

a. Reported in Section 3.3
3. How do these results compare with reporting in published articles, specifically:

a. Are there discrepancies in the number of reported volunteers challenged?

i. Reported in Section 3.4 B
b. Are there discrepancies in the number of reported volunteers positive for infection?

i. Reported in Section 3.4 C
c. Are there discrepancies in AE reporting?

i. Reported in Section 3.4 D
d. Are there discrepancies in SAE reporting?

i. Reported in Section 3.4 E
e. Are there discrepancies in the level of detail of reported adverse events?

i. Reported in Section 3.4 F
4. Based on the quality of data reporting and record metadata (such as length of trials and number of trials that are not completed), do CHIS make effective use of ClinicalTrials.gov?

a. Reported in Section 3.4 F

### Search Strategy

A query was performed on 4/15/23 using the AACT API with the following terms:

~~~
select * from studies where
study_type = ‘Interventional’
and
enrollment < 1000
and
study_first_submitted_date < ‘2022-06-30’
and
(
(official_title ilike ‘%challenge%’) or
(official_title ilike ‘%immunization%’ and official_title ilike ‘%sporozoites%’) or
(official_title ilike ‘%human%’ and official_title ilike ‘%carriage%’) or
(official_title ilike ‘%infection%’ and
(official_title ilike ‘%controlled%’ or official_title ilike ‘%experimental%’ or official_title ilike ‘%induced%’)) or
(official_title ilike ‘%efficacy%’ and official_title ilike ‘%vaccine%’) or
(official_title ilike ‘%human%’ and official_title ilike ‘%exposure%’) or
(official_title ilike ‘%healthy%’ and
(official_title ilike ‘%naïve%’ or official_title ilike ‘%naive%’)) or
(official_title ilike ‘%competitive%’ and official_title ilike ‘%carriage%’)
OR
(brief_title ilike ‘%challenge%’) or
(brief_title ilike ‘%immunization%’ and brief_title ilike ‘%sporozoites%’) or
(brief_title ilike ‘%human%’ and brief_title ilike ‘%carriage%’) or
(brief_title ilike ‘%infection%’ and
(brief_title ilike ‘%controlled%’ or brief_title ilike ‘%experimental%’ or brief_title ilike ‘%induced%’)) or
(brief_title ilike ‘%efficacy%’ and brief_title ilike ‘%vaccine%’) or
(brief_title ilike ‘%human%’ and brief_title ilike ‘%exposure%’) or
(brief_title ilike ‘%healthy%’ and
(brief_title ilike ‘%naïve%’ or brief_title ilike ‘%naive%’)) or
(brief_title ilike ‘%competitive%’ and brief_title ilike ‘%carriage%’)
OR
(acronym ilike ‘%challenge%’) or
(acronym ilike ‘%human%’)
OR
nct_id IN
(select s.nct_id from studies s, keywords k where
s.nct_id = k.nct_id and k.name ilike ‘%challenge%’)
OR
nct_id IN
(select s.nct_id from studies s, detailed_descriptions d where
s.nct_id = d.nct_id and
((d.description ilike ‘%challenge%’) and
(d.description ilike ‘%infection%’ or
d.description ilike ‘%controlled%’ or
d.description ilike ‘%experimental%’)))
OR
nct_id IN
(select s.nct_id from studies s, brief_summaries b where
s.nct_id = b.nct_id and
((b.description ilike ‘%challenge%’) and
(b.description ilike ‘%infection%’ or
b.description ilike ‘%controlled%’ or
b.description ilike ‘%experimental%’)))
)
~~~

For record identified by this query, an additional query was performed to aggregate AE data:

~~~
select cv.nct_id, cv.number_of_nsae_subjects, cv.minimum_age_num, cv.maximum_age_num,
 dg.design_groups,
 iv.interventions,
 oap.p_value,oac.ci_percent,
 srp.pmid,src.citation,
 pf.recruitment_details,
 rd.AE_Count,rd.SAE_Count,rd.Mortality_Count,
 re.Num_AEs_described
~~~

~~~
 from (
 select calculated_values.nct_id, calculated_values.number_of_nsae_subjects,
calculated_values.minimum_age_num, calculated_values.maximum_age_num
 from calculated_values
 where calculated_values.nct_id = ‘{}’) as cv
 left join (
 select design_groups.nct_id, string_agg(design_groups.description,’; ‘) as design_groups
 from design_groups
 group by design_groups.nct_id) as dg
 on cv.nct_id = dg.nct_id
 left join (
 select interventions.nct_id, string_agg(interventions.description,’;’) as interventions
 from interventions
 group by interventions.nct_id) as iv
 on cv.nct_id = iv.nct_id
 left join (
 select outcome_analyses.nct_id, string_agg(CAST(outcome_analyses.p_value as VarChar),’;
’) as p_value
 from outcome_analyses
 group by outcome_analyses.nct_id) as oap
 on cv.nct_id = oap.nct_id
 left join (
 select outcome_analyses.nct_id, string_agg(CAST(outcome_analyses.ci_percent as
VarChar),’; ‘) as ci_percent
 from outcome_analyses
 group by outcome_analyses.nct_id) as oac
 on cv.nct_id = oac.nct_id
 left join (
 select study_references.nct_id, string_agg(CAST(study_references.pmid as VarChar),’; ‘) as
pmid
 from study_references
 group by study_references.nct_id) as srp
 on cv.nct_id = srp.nct_id
 left join (
 select study_references.nct_id, string_agg(CAST(study_references.citation as VarChar),’; ‘)
as citation
 from study_references
 group by study_references.nct_id) as src
 on cv.nct_id = src.nct_id
 left join (
 select participant_flows.nct_id, string_agg(CAST(participant_flows.recruitment_details as
VarChar),’; ‘) as recruitment_details
 from participant_flows
 group by participant_flows.nct_id) as pf
 on cv.nct_id = pf.nct_id
 left join (
 select reported_events.nct_id, COUNT(DISTINCT reported_events.adverse_event_term) AS
Num_AEs_described
 from reported_events
 group by reported_events.nct_id) as re
 on cv.nct_id = re.nct_id
~~~

~~~
 left join(
 select reported_event_totals.nct_id,
 sum(case when reported_event_totals.classification = ‘Total, other adverse events’ then
 reported_event_totals.subjects_affected else 0 end) as AE_Count,
 sum(case when reported_event_totals.classification = ‘Total, serious adverse events’ then
 reported_event_totals.subjects_affected else 0 end) as SAE_Count,
 sum(case when reported_event_totals.classification = ‘Total, all-cause mortality’ then
 reported_event_totals.subjects_affected else 0 end) as Mortality_Count
 from reported_event_totals
 group by reported_event_totals.nct_id) as rd
 on cv.nct_id = rd.nct_id
~~~

This search returned 5,131 results. A filter by study size was applied using a cohort size of 1,000 as an upper limit. This is based on our previous systematic review, in which the largest CHIS identified had a cohort size of 437. This number was rounded to 500 and doubled to 1000 to ensure a conservative ceiling on sample size to maximize the number of eligible CHIS.

Interventional studies were used as filter criteria because CHIS are rarely classified as another type of study.

### Systematic review protocol

Titles and brief descriptions for 5,131 results (ClinicalTrials.gov records) were screened for inclusion by 2 of 6 reviewers. In instances in which the title or brief description did not provide sufficient evidence for challenge, the arms of the trials were checked for challenge phases. Records that described a study that intentionally exposed human participants to an infectious pathogen for the purpose of modeling a named human disease were included. Records that did not intentionally expose a human cohort to an infectious pathogen (such as studies involving a live vaccine that was not given with intent to challenge, studies using non-infectious challenge agents, and studies that performed noxious challenges with lipopolysaccharide or endotoxin) were excluded.

### Eligibility criteria

Clinical trial records will be reviewed for the following criteria:

1) Trial intentionally exposes at least one volunteer to an infectious pathogen

Clinical trial records will be excluded for the following criteria:

1) Trial record is not in english
2) Trial record does not have sufficient information to determine the procedure and primary outcomes of the trial
3) Trial does not intentionally expose at least one volunteer to an infectious pathogen.

Trials that expose volunteers to live-attenuated pathogens will be included, unless the agent used is explicitly attenuated to the degree of being entirely non-pathogenic. Non-pathogenic agents often used in challenge studies, such as lipopolysaccharide or gluten, will not be included. In the current definition, an infectious agent must be a biological agent with the capacity to reproduce and cause infection in its wild-type, unattenuated form.

### Methods for identifying full-text articles

Because ClinicalTrials.gov lists publications of trial results as part of the record, full-text articles will be identified from the list given in the record. All publications listed in the ClinicalTrials.gov record will be evaluated. A PubMed search for the trial’s NCT number will be performed to identify publications not listed in the ClinicalTrials.gov record.

### Statistical analysis plan

Trials that have been completed or are currently ongoing will be included in data analysis. Trials that have not yet begun, have been withdrawn, or were completed within 1 year of the date data collection was completed (4/15/23) will be included but not analyzed.

The unit of analysis will be the rate of discrepancy between reporting in ClinicalTrials.gov records and associated publications. Data will be divided into groups by sponsor type (private, public, and public-private partnership), study size (1st, 2nd, 3rd, and 4th quartiles), and risk of bias (low, some concerns, high).

Odds ratios and 95% confidence intervals will be used to evaluate significance between the rates of discrepancy with each subgroup compared to the remainder of its group. These analyzes will be performed post-hoc and were not pre-registered.

### Data items

Reported AE’s will be assessed for the following qualities:

a) reporting of frequency of AE assessment,
b) duration of follow up for AEs,
c) investigators attribution of the cause of SAEs, categorized as related or unrelated to challenge,
d) what proportion of AEs determined by the investigators to be unexpected

The following data will be automatically extracted from CT records:

- Status of trial (Active but not recruiting, Completed, Enrolling by invitation, Not yet recruiting, Terminated, Unknown status, or Withdrawn)
- The date the study was posted
- The date results were posted posted
- Study type
- Enrollment
- Minimum age
- Maximum age
- Number of volunteers with at least 1 SAE
- Number of volunteers with at least 1 AE
- All cause mortality
- Number of potential AEs described
- Study sponsor

The following data will be manually extracted from CT records:

- Number of volunteers in challenge groups
- Number of volunteers in challenge groups that became infected with the pathogen
- Are AEs detailed individually? (true/false)
- Total number of publications
- Number of publications automatically indexed
- Number of volunteers with AEs prior to challenge
- Number of volunteers with AEs after challenge
- Number of volunteers with AEs after rechallenge

The following data will be manually extracted from associated publications:

- Study enrollment
- Number of volunteers in challenge groups
- Number of volunteers in challenge groups that became infected with the pathogen
- Number of volunteers with at least 1 SAE
- Number of volunteers with at least 1 AE
- All cause mortality
- Are AEs detailed individually? (true/false)
- Number of volunteers with AEs prior to challenge
- Number of volunteers with AEs after challenge
- Number of volunteers with AEs after rechallenge

Automatic data extraction will be reviewed by 2 of 6 reviewers. Manual data extraction for each included record and any publications associated with the record that discuss results of the study will also be performed by 2 of 6 reviewers. Conflicting data reported by different reviewers will be resolved by DT or JAP.

### Risk of bias in individual records

Risk of bias in individual records will be assessed using the Cochrane Risk of Bias 2.0 Tool by 2 of 6 reviewers. The Cochrane Risk of Bias 2.0 tool evaluates bais using separate algorithms to rate the risk of bias from 5 potential sources in publications. In the present study, bias arising from Domain 5, “Risk of bias in the selection of the reported result,” will be evaluated for ClinicalTrials.gov records and all associated publications. Results of these assessments for ClinicalTrials.gov records and associated publications will be combined for a single assessment given per ClinicalTrial.gov record. Disputes will be resolved by JAP or DT.

### Data synthesis

ClinicalTrials.gov records will be determined eligible for analysis if they post results and have an associated publication linked to their NCT number. Data will be tabulated to create summary statistics by relevant parameters as detailed in the analysis grouping below.

Analysis grouping:

- Trial recruitment status

- Completed: The primary group used for analyzing data
- Active, not recruiting: Used to analyze the number of current studies (and when they started, how long they’ve been in progress, estimated completion, etc.)
- Recruiting (and Enrolling by invitation): Used to analyze the number of current studies (and when they started, how long they’ve been in progress, estimated completion, etc.)
- Withdrawn (and Terminated): Used to analyze studies that were proposed (or even started) that were not finished, in order to identify and investigate any studies that exposed participants to a challenge agent, but did not finish the study or report results of the challenge
- Unknown status: Studies that haven’t been updated within a certain timeframe; will be analyzed individually for available data
- Sponsor type

- Private sponsorship: Private for-profit or not-for-profit organizations
- Public sponsorship: Governmental organizations
- Public-private partnership: An independent collaborative partnership between a governmental organization and a private organization
- Study size

- 1st Quartile: The 1st quartile of enrollment size for all studies included in analysis
- 2nd Quartile: The 2nd quartile of enrollment size for all studies included in analysis
- 3rd Quartile: The 3rd quartile of enrollment size for all studies included in analysis
- 4th Quartile: The 4th quartile of enrollment size for all studies included in analysis
- Risk of bias

- Low risk of bias: A low risk of bias as determined by the algorithm provided Domain 5 of the Cochrane Risk of Bias 2.0 Tool, taking into account results reported in the ClinicalTrials.gov record and all publications associated with its NCT number.
- Some concerns: Some concerns as determined by the algorithm provided Domain 5 of the Cochrane Risk of Bias 2.0 Tool, taking into account results reported in the ClinicalTrials.gov record and all publications associated with its NCT number.
- High risk of bias: A high risk of bias as determined by the algorithm provided Domain 5 of the Cochrane Risk of Bias 2.0 Tool, taking into account results reported in the ClinicalTrials.gov record and all publications associated with its NCT number.

## Footnotes

1 A range of values was given to account for unclear data reporting in some studies.

